# Genetic Overlap Between Inflammatory Bowel Disease and Neurological Disorders: Insights from GWAS and Gene Expression Analysis

**DOI:** 10.1101/2024.07.29.24311160

**Authors:** Utkarsh Tripathi, Yam Stern, Inbal Dagan, Ritu Nayak, Eva Romanovsky, Eran Zittan, Shani Stern

**Affiliations:** Sagol Department of Neurobiology, Faculty of Natural Sciences, University of Haifa, Israel; The Rappaport Faculty of Medicine Technion-Israel Institute of Technology, Haifa, Israel

## Abstract

Inflammatory Bowel Disease (IBD), which includes Crohn’s disease (CD) and Ulcerative Colitis (UC), is a complex and multifactorial condition marked by chronic inflammation of the gastrointestinal tract. This study leverages genome-wide association studies (GWAS) and gene expression data from the Genotype-Tissue Expression (GTEx) project to investigate IBD’s genetic and expression profiles and subtypes. We examined 207 studies related to IBD, 71 specific to CD, and 66 focused on UC, identifying both shared and unique genetic factors among these conditions. GWAS meta-analysis revealed the top IBD-associated genes, which include IL23R, NOD2, ATG16L1, HLA-DRB9, and more. Pathway enrichment analyses identified consistently enriched pathways such as the NF-kappa B signaling pathway, JAK-STAT signaling pathway, and cytokine-cytokine receptor interaction, all of which play critical roles in immune responses and inflammation. Gene Ontology (GO) term analysis highlighted processes like cytokine production, cell activation, and leukocyte activation, reinforcing their involvement in the pathogenesis of IBD. Gene expression analysis showed that genes associated with IBD are expressed not only in the gastrointestinal tract but also in various regions of the brain, suggesting potential links between IBD and neurological functions. Our study further explored the genetic overlap between IBD and several neurological disorders, including schizophrenia, depression, autism spectrum disorder, and attention-deficit/hyperactivity disorder, uncovering a shared genetic architecture. These findings emphasize the systemic nature of IBD and its potential neurological implications, paving the way for targeted therapeutic strategies that address both gastrointestinal and neurological aspects of the disease.

## Introduction

Inflammatory Bowel Disease (IBD) is a chronic, relapsing inflammatory condition of the gastrointestinal tract, primarily encompassing Crohn’s Disease (CD) and Ulcerative Colitis (UC). Despite extensive research, the precise etiology of IBD remains elusive, though it is widely accepted to be a multifactorial disorder influenced by genetic, environmental, and immunological factors (Franke A. et al., 2010; Jostins L et al., 2012). The identification of genetic loci associated with IBD through genome-wide association studies (GWAS) has significantly advanced our understanding of the disease, yet much remains to be uncovered about the functional implications of these genetic variations (Xavier R, Podolsky D, 2007).

CD and UC, the two primary forms of IBD, present distinct clinical and pathological features, yet they share overlapping genetic and environmental risk factors. CD can affect any part of the gastrointestinal tract from mouth to anus, often leading to transmural inflammation, while UC is typically confined to the colon and rectum, with inflammation limited to the mucosal layer (Daneese S, Fiocchi C, 2011). Despite these differences, both conditions are characterized by chronic inflammation, mucosal damage, and an increased risk of colorectal cancer (Baumgart D, Carding S, 2007).

Recent advances in high-throughput genomic technologies have enabled the comprehensive analysis of genetic and transcriptomic data, facilitating the identification of IBD-associated genes and their expression patterns across different tissues (Lonsdale et al., 2013). The integration of data from resources such as the Genotype-Tissue Expression (GTEx) project has further allowed for the exploration of gene expression in various tissues, including those outside the gastrointestinal tract, such as the brain (Carithers L, Moore H, 2015). This holistic approach provides valuable insights into the systemic nature of IBD and its potential impact on extra-intestinal tissues (Liu J, van Sommeren S, 2015).

The brain-gut axis is the complex communication network linking the central nervous and gastrointestinal systems. The gut affects the brain through various mechanisms, notably the production of neurotransmitters and hormones. For example, the gut microbiota produces short-chain fatty acids that can influence the brain’s function and mood regulation (Cryan J, Dinan T, 2012). Additionally, gut receptors can signal the brain about hunger and satiety, impacting emotional states and cognitive processes. This relationship highlights the gut’s role as a vital component in mental health and cognition, indicating that gut health can directly influence emotional well-being.

Conversely, the brain can also significantly affect gut function through the autonomic nervous system. Stress and emotional disturbances can lead to intestinal motility and permeability changes, often leading to gastrointestinal symptoms. When the brain perceives stress, it can trigger the release of stress hormones such as cortisol, which may disrupt the gut’s normal function. This communication pathway means that psychological states can manifest as physical symptoms in the gut, further underscoring the dynamic relationship between these two systems.

Mental stress is particularly relevant in the context of inflammatory bowel disease (IBD), as it can exacerbate flares and increase symptoms. Stressful events may lead to an imbalance in the gut microbiome and heightened inflammatory responses, which are critical factors in IBD. Many individuals with IBD report that high-stress moments correlate with worsening symptoms, such as abdominal pain and diarrhea. Managing stress through therapeutic interventions or lifestyle changes can, therefore, be an essential aspect of treating IBD, highlighting the importance of considering mental health in the overall management of gut-related disorders.

Moreover, the overlap between IBD and various neurological and psychiatric disorders, including schizophrenia, depression, autism spectrum disorder (ASD), and attention-deficit/hyperactivity disorder (ADHD), suggests shared genetic and immunological pathways (Benros M et al., 2013; Mikocka-Walus A et al., 2007; Bhamre R et al., 2018, Bernstein C et al., 2019). Studies show that depression and anxiety are more prevalent in individuals with IBD. Some also suggest a potential association with Schizophrenia (Sung K et al., 2022). These comorbidities underscore the importance of understanding the genetic underpinnings of IBD not only within the context of gastrointestinal pathology but also concerning broader systemic and neurological health.

This review aims to synthesize the current understanding of the genetic landscape of IBD, CD, and UC by leveraging GWAS data and integrating findings from the GTEx project. We will explore the expression patterns of IBD-associated genes across different tissues, mainly focusing on the gastrointestinal tract and brain, and highlight the enriched biological pathways. Furthermore, we will examine the genetic overlap between IBD and several brain disorders, providing a comprehensive view of the shared genetic architecture and potential common pathways. This integrated analysis aims to advance our understanding of IBD pathogenesis and its broader systemic implications, paving the way for targeted therapeutic strategies and improved patient outcomes.

## Methods

### Data Collection and Preprocessing

#### GWAS Data

Genome-wide association studies (GWAS) data for IBD, CD, and UC were obtained from publicly available databases. The IBD dataset comprises 2700 associations from 207 studies, the CD dataset comprises 1092 associations from 71 studies, and the UC dataset comprises 801 associations from 66 studies, providing a comprehensive overview of genetic variants linked to IBD and its subtypes. To ensure consistency in gene nomenclature across the GWAS data, the Human Gene Nomenclature Committee (HGNC) gene symbols were standardized using the HGNC database, which contains approved gene symbols, previous symbols, and aliases linked to Ensemble IDs. Following protocols described in previous studies (Choudhary A et al., 2022; Romanovsky E et al., 2023), this standardization ensures accurate identification and comparison of genetic data across different datasets.

#### GTEx Data

Expression data for IBD-associated genes were obtained from the Genotype-Tissue Expression (GTEx) project. The GTEx dataset provides comprehensive gene expression profiles across multiple human tissues. We specifically extracted expression data for gastrointestinal (GIT) tract tissues and various brain regions. The data included an Ensemble gene identifier (Gene), analyzed sample (Tissue), and normalized expression value (TPM) for each gene. TPM values are a normalization method for RNA-Seq data, accounting for the gene length and sequencing depth, allowing for accurate comparisons of gene expression levels across samples. We had expression data for the following gastrointestinal (GIT) areas: Colon – Sigmoid, Esophagus – Gastroesophageal Junction, Esophagus – Mucosa, Esophagus – Muscularis, Stomach, Colon – Transverse, and Small Intestine – Terminal Ileum. For brain regions, we had data from: the Amygdala, Anterior Cingulate Cortex (BA24), Caudate (basal ganglia), Cerebellar Hemisphere, Cerebellum, Cortex, Frontal Cortex (BA9), Hippocampus, Hypothalamus, Nucleus Accumbens (basal ganglia), Putamen (basal ganglia), Spinal Cord (cervical C1), and Substantia Nigra.

#### Pathway Enrichment Analysis

Pathway enrichment analysis was performed to identify biological pathways significantly enriched with IBD-associated genes. The Kyoto Encyclopedia of Genes and Genomes (KEGG) and Gene Ontology (GO) databases were used for pathway annotations. Enrichment analysis was conducted using the Shiny-GO tool. Pathways with a false discovery rate (FDR) adjusted p-value < 0.05 were considered significantly enriched.

#### Brain and Signaling-Related Pathway Analysis

To explore the potential neurological implications of IBD-associated genes, we focused on brain-related pathways. We were specifically interested in KEGG pathways, which were directly related to brain or signaling, and these were plotted in addition to the general KEGG pathways. The identified pathways were visualized using bar plots to illustrate the fold enrichment and gene ratio for each pathway.

#### Single Nucleotide Polymorphism (SNP) Analysis

The SNP data for UC and CD were obtained from the GWAS Catalog. To identify common genes, we first extracted genes associated with UC and CD from the GWAS data. The intersection of genes from UC and CD GWAS datasets was then determined. Unique SNPs from both datasets were combined and mapped to these common genes.

A matrix was constructed to represent the presence of SNPs in common genes, with each entry indicating whether the SNP was associated with UC, CD, or both. The total number of SNPs per gene was calculated by summing the non-empty entries in each row of the matrix. The SNP percentage for each gene was then calculated by dividing the total number of SNPs for that gene by the total number of SNPs across all genes and multiplying by 100.

Genes were filtered based on SNP percentage, retaining those with more than 1% of the total SNPs to focus on genes significantly associated with the disease. The oncoPrint function from the ComplexHeatmap R package was used to generate a heatmap, with colors indicating the association of SNPs with UC, CD, or both. This visual representation highlighted the overlap and distinctions in SNP associations between UC and CD, providing insights into conditions’ genetic commonalities and differences.

#### Word Cloud Generation

To visualize the prominence of genes associated with Inflammatory Bowel Disease (IBD), UC, and CD, word clouds were generated using Python3.0. The word clouds highlight each gene’s frequency by presenting the gene’s sizn proportionality to the frequency it was reported in the literature, providing a visual representation of the most commonly studied genes in each condition.

### Genetic Overlap Analysis

#### Overlap with Brain Disorders

We investigated the genetic overlap between IBD and several brain disorders, including schizophrenia (SCZ), depression, autism spectrum disorder (ASD), and attention-deficit/hyperactivity disorder (ADHD). Gene lists for these disorders were obtained from relevant GWAS studies and databases. Venn diagrams were generated to visualize the overlap between IBD and each brain disorder, highlighting common genes.

#### Overlap Among IBD Subtypes

The overlap between genes associated with IBD, CD, and UC was examined. Venn diagrams were used to illustrate the shared and unique genes among these subtypes. Additionally, an UpSet plot was generated to provide a detailed view of the intersections between IBD subtypes and brain disorders, allowing for the identification of genes common to multiple conditions.

### Gene Expression Analysis

#### Average Gene Expression Calculation

For IBD-associated genes, the average expression levels across different GIT and brain tissues were calculated. This involved computing each tissue type’s mean TPM values for IBD genes. The resulting expression profiles were visualized using heat maps to highlight the distribution of IBD gene expression in the different tissues.

Biorender.com was utilized to assign color-coding based on the ranking of average expression level in the brain and GIT, with red signifying a high expression level and violet representing a low expression level. The illustration is not a precise representation of the anatomical structure of the brain and GIT but rather a schematic one. The colored areas were used to demonstrate the different brain and GIT regions.

#### Visualization

All visualizations, including heatmaps, bar plots, Venn diagrams, and UpSet plots, were generated using the R programming language and Python 3.0 and relevant packages (e.g., ggplot2, ComplexHeatmap, UpSetR). The visualizations were designed to provide clear and comprehensive representations of the data, facilitating the interpretation of gene expression patterns and pathway enrichment results.

## Results

### Genetic Associations in Inflammatory Bowel Disease (IBD)

The genetic architecture of Inflammatory Bowel Disease (IBD) encompasses a wide array of genes that contribute to the complex etiology of the disease. Utilizing GWAS data, we identified a comprehensive list of genes associated with IBD. As shown in Figure 1a, the word cloud highlights the most frequently associated genes with IBD, with the size of each gene’s name reflecting its publication frequency (see Methods). Notably, IL23R is the most prominent, appearing in 38 publications, followed by NOD2 (22 publications), PUS10 (15 publications), IRF1 (15 publications), ATG16L1 (15 publications), and C1orf141 (15 publications). These genes are known to be involved in immune regulation and inflammatory responses, which are central to the pathogenesis of IBD (Franke A. et al., 2010; Jostins L. et al., 2012). We further conducted pathway enrichment analyses to explore these genetic associations’ functional implications. Figure 1b presents the top 10 KEGG pathways that are enriched with IBD-associated genes. The most significantly enriched pathways include the Inflammatory Bowel Disease (13 genes, FDR =2.8E-09, nFold = 12.74) (IBD), Th17 Cell Differentiation(15 genes, FDR = 1.3E-08, nFold = 8.84), NF-Kappa B Signaling Pathway(13 genes, FDR = 4.0E-07, nFold = 7.96), Cytokine-Cytokine Receptor Interaction(21 genes, FDR = 4.0E-07, nFold = 4.55), and Th1 and Th2 Cell Differentiation(12 genes, FDR = 7.9E-07, nFold = 8.31). These pathways are crucial in mediating immune and inflammatory responses. The enrichment of the Th17 Cell Differentiation pathway is particularly noteworthy, given its role in driving chronic inflammation in IBD (Baumgart D, Carding S, 2007; Kaser A, Zeissig S, Blumberg R, 2010). The NF-Kappa B Signaling Pathway is a key regulator of immune response and inflammation, playing a pivotal role in the transcription of pro-inflammatory cytokines (Atreya I, Atreya R, Neurath M, 2008). The Cytokine-Cytokine Receptor Interaction pathway, which involves various cytokines and their receptors, is essential for cell signaling in immune responses, highlighting the importance of cytokine networks in the pathology of IBD (Chen M, Sundrud M, 2016; Sanchez-Munoz F et al., 2008). Additionally, pathways such as Th1 and Th2 Cell Differentiation are involved in the adaptive immune response, with Th1 cells being critical for mediating responses against intracellular pathogens and Th2 cells being important for humoral immunity and allergic responses (Chen M, Sundrud M, 2016; Imam T et al., 2018).

**Figure 1:** Genetic Associations in Inflammatory Bowel Disease (IBD) **(a)** A word cloud of genes associated with IBD. The size of each gene’s name is proportional to the frequency of its mention in the literature. Prominent genes include NOD2, IL23R, C1orf141, ATG16L1, and PUS10. **(b)** The top 10 enriched KEGG pathways in IBD-associated genes. Significant pathways include the Inflammatory Bowel Disease Pathway, Th17 Cell Differentiation, and NF-Kappa B Signaling Pathway. **(c)** The top 10 enriched GO Biological Process terms in IBD GWAS data. Enriched terms include Leukocyte Activation, Regulation of Cytokine Production, and Positive Regulation of Multicellular Organismal Processes.

Figure 1c shows the top 10 Gene Ontology (GO) Biological Process terms enriched in the IBD GWAS data. The most enriched terms include Leukocyte Activation (73 genes, FDR = 1.3E-15, nFold = 3.15), Regulation of Cytokine Production (59 genes, FDR = 9.7E-17, nFold = 4.04), Positive Regulation of Multicellular Organismal Process (77 genes, FDR = 1.3E-15, nFold = 3.04), Lymphocyte Activation (53 genes, FDR = 2.3E-15, nFold = 4.09), and Positive Regulation of Immune System Process (61 genes, FDR = 1.2E-13, nFold = 3.26). These biological processes are essential for the initiation and maintenance of immune responses. For instance, the regulation of cytokine production involves a network of signaling molecules that mediate and regulate immunity, inflammation, and hematopoiesis, which are pivotal in the inflammatory milieu of IBD (Neurath M, 2014).

The consistent enrichment of terms related to cytokine production and immune system processes suggests a vital role of these processes in IBD pathophysiology. The term Leukocyte Activation involves the activation of white blood cells, which play a crucial role in the immune response by identifying and eliminating pathogens (Enkai S et al., 2003). Similarly, Lymphocyte Activation is critical for the adaptive immune response, involving the activation of T cells and B cells, which are essential for targeted immune responses (Kanakasabai S et al., 2010).

### Genetic Associations in CD (IBD)

To delve deeper into the genetic underpinnings specific to CD, we examined a separate subset of the GWAS data that is specific to CD. Figure 2a shows a word cloud of genes associated with CD. Prominent genes include IL23R, NOD2, ATG16L1, C1orf141, and IRF1-AS1, consistent with those identified in the broader IBD analysis, underscoring their critical roles in the disease. Notably, IL23R appears in 21 publications, making it the most significant gene, followed by NOD2 (18 publications), ATG16L1 (13 publications), C1orf141 (12 publications), and IRF1-AS1 (10 publications), reinforcing their central roles in CD pathogenesis (Naser S et al., 2012).

**Figure 2:** Genetic Associations in CD. **(a)** A word cloud of genes associated with CD. The size of each gene’s name reflects its frequency of mention in the literature. Prominent genes include NOD2, IL23R, C1orf141, ATG16L1, and IRF1-AS1. **(b)** The top 10 enriched KEGG pathways in CD-associated genes. Significant pathways include the Inflammatory Bowel Disease Pathway, Th17 Cell Differentiation, and JAK-STAT Signaling Pathway. **(c)** The top 10 enriched GO Biological Process terms in CD GWAS data. Enriched terms include inflammatory response, Cytokine Production, and Positive Regulation of signal transduction.

The pathway enrichment analyses for CD, as depicted in Figure 2b, reveal significant insights into the functional implications of these genetic associations. hed pathways include the Inflammatory Bowel Disease(12 genes, FDR= 1.5E-09, nFold=16.7), Th17 Cell Differentiation(14 genes, FDR= 1.9E-09, nFold=11.73), NF-Kappa B Signaling Pathway(11 genes, FDR= 1.2E-06, nFold=9.57), Cytokine-Cytokine Receptor Interaction (17 genes, FDR= 1.2E-06, nFold=5.23), and Th1 and Th2 Cell Differentiation (10 genes, FDR= 2.0E-06, nFold=9.83). The JAK-STAT Signaling Pathway (13 genes, FDR= 1.2E-06, nFold=7.26) is also prominently enriched, highlighting its critical role in transmitting extracellular signals to the cell nucleus, influencing gene expression and immune responses (Schindler C et al., 2007). This pathway’s significance in CD is further supported by its involvement in various cytokine-signaling mechanisms that regulate immune function.

Figure 2c illustrates the top 10 Gene Ontology (GO) Biological Process (BP) terms enriched in CD, highlighting the role of immune and inflammatory responses. Key processes include “Cytokine Production” (49 genes, FDR = 1.5E-16, nFold = 4.73), “Regulation of Cytokine Production” (49 genes, FDR = 1.5E-16, nFold = 4.77), and “Leukocyte Activation” (59 genes, FDR = 3.8E-15, nFold = 3.62), which are crucial for understanding the disease’s immune-related mechanisms. The terms “Cell Activation” (62 genes, FDR = 8.2E-15, nFold = 3.38) and “Positive Regulation of Multicellular Organismal Process” (61 genes, FDR = 8.2E-15, nFold = 3.42) further emphasize the involvement of systemic immune regulation in CD. Additionally, “Lymphocyte Activation” (42 genes, FDR = 7.1E-14, nFold = 4.61) and “Inflammatory Response” (43 genes, FDR = 1.3E-12, nFold = 4.09) highlight the importance of immune cells in maintaining chronic inflammation in the disease. These processes provide insights into the underlying biological mechanisms of CD and potential therapeutic targets (Baharvand H et al., 2007).

The enrichment of BP terms like Cytokine Production and Regulation of Cytokine Production suggest a possible central role of cytokine networks in CD pathophysiology. Such networks facilitate cell communication during immune responses (Alhendi A, Naser S, 2023; Vebr M et al., 2023).

These findings provide a detailed understanding of the genetic and molecular landscape specific to CD, emphasizing the significant overlap in genetic and pathway associations with the broader IBD category while highlighting specific processes and signaling pathways that contribute uniquely to CD pathology.

### Genetic Associations in Ulcerative Colitis

Continuing from the genetic insights gained from the broader IBD and CD analyses, we now focus on UC. Figure 3a displays a word cloud of genes associated with UC, with the size of each gene’s name reflecting its frequency in the GWAS data. Prominent genes include IL23R (17 publications), IFNG-AS1 (9 publications), HLA-DRB9 (8 publications), INAVA (7 publications), CARD9 (7 publications), and LINC01620 (7 publications). Notably, IL23R is common across IBD (38 publications), CD (21 publications), and UC, underscoring its critical role in these conditions (Duerr R et al., 2006; Peng L et al., 2017; Sewell G, Kaser L, 2022). Additionally, NOD2 and ATG16L1, which are highly prominent in IBD (22 and 15 publications, respectively) and CD (18 and 13 publications, respectively), were observed in only 4 and 3 publications in UC, respectively, indicating a lesser but still notable involvement in UC pathogenesis.

**Figure 3:** Genetic Associations in Ulcerative Colitis. **(a)** A word cloud of genes associated with UC. The size of each gene’s name reflects its frequency of mention in the literature. Prominent genes include IL23R, HLA-DRB9, CARD9, IFNG-AS1, and INAVA. **(b)** The top 10 enriched KEGG pathways in UC-associated genes. Significant pathways include the HIF-1 Signaling Pathway, TNF Signaling Pathway, and JAK-STAT Signaling Pathway. **(c)** The top 10 enriched GO Biological Process terms in UC GWAS data. Enriched terms include Cell Activation, Cytokine Production, and Regulation of Cytokine Production.

The pathway enrichment analyses for UC, illustrated in Figure 3b, reveal significant insights into the functional implications of these genetic associations. s, FDR= 1.2E-05, nFold=9.5), TNF Signaling Pathway (9 genes, FDR= 1.2E-05, nFold=9.25), JAK-STAT Signaling Pathway(11 genes, FDR= 5.9E-06, nFold=7.8), NF-Kappa B Signaling Pathway (10 genes, FDR= 9.3E-07, nFold=11.07), and Cytokine-Cytokine Receptor Interaction (16 genes, FDR= 3.0E-07, nFold=16). These pathways highlight both common and unique aspects of UC pathogenesis compared to CD and the broader IBD category.

The HIF-1 Signaling Pathway is noteworthy for its role in the cellular response to hypoxia, which can influence inflammation and tissue remodeling in UC (Holmqvist E et al., 2010). This pathway is also enriched in CD and the broader IBD category, indicating its significance across these conditions. The TNF Signaling Pathway, known for mediating inflammatory processes, is a common feature in both UC and CD, reflecting its pivotal role in IBD pathology (Choi S et al., 2000). The JAK-STAT Signaling Pathway is crucial for transducing signals from various cytokines and growth factors, influencing gene expression and cellular function. This pathway is significantly enriched in both UC and CD, highlighting its importance in immune regulation across IBD subtypes (Coskun M et al., 2013; Schindler C et al., 2007).

The NF-Kappa B Signaling Pathway and Cytokine-Cytokine Receptor Interaction pathways are also shared between UC and CD, emphasizing their central role in regulating immune responses and inflammation in both conditions (Atreya I, Atreya R, Neurath M, 2008; Vebr M et al., 2023). These pathways are integral to the broader IBD pathology, underscoring the shared mechanisms underlying these diseases.

Figure 3c presents the top 10 Gene Ontology (GO) Biological Process terms enriched in the UC GWAS data. Key processes include Cell Activation (53 genes, FDR = 3.6E-14, nFold = 3.68), Cytokine Production (40 genes, FDR = 3.6E-14, nFold = 4.91), Regulation of Cytokine Production (40 genes, FDR = 3.6E-14, nFold = 4.95), Leukocyte Activation (50 genes, FDR = 3.6E-14, nFold = 3.90), and Inflammatory Response (39 genes, FDR = 2.9E-13, nFold = 4.73). These biological processes are critical for initiating and maintaining immune responses, with cytokine production and regulation being pivotal in the inflammatory environment of UC (Xavier R, Podolsky D, 2007).

The enrichment of Lymphocyte Activation (38 genes, FDR = 3.6E-14, nFold = 5.30) is shared across UC, CD, and the broader IBD category, underscoring the role of adaptive immune responses in these conditions. Lymphocytes, including T and B cells, are essential for targeted immune responses and play a significant role in the pathogenesis of UC (Kałużna A et al., 2022). The Positive Regulation of Multicellular Organismal Process (50 genes, FDR = 7.9E-13, nFold = 3.57) and Regulation of Response to External Stimulus (42 genes, FDR = 3.3E-11, nFold = 3.77) terms further highlight the complex interactions between immune cells and their environment in UC.

The Negative Regulation of Response to Stimulus (32 genes) and T Cell Activation (28 genes) terms reflect the intricate balance of immune activation and regulation required to maintain homeostasis and prevent excessive inflammation in UC. These terms are also enriched in CD, indicating shared regulatory mechanisms in these IBD subtypes.

### IBD genes and their expression in the gut and brain

Building upon the genetic insights from IBD, CD, and UC, we utilized GTEx data to examine the expression of IBD-associated genes across different tissues. We had expression data for seven specific gastrointestinal (GIT) areas and 15 brain areas (mentioned in Methods). Figure 4a illustrates the average expression levels of these genes in various regions of the gastrointestinal (GIT) tract. High expression levels are observed in the esophagus muscularis, gastroesophageal junction, terminal ileum, and transverse colon, among others. This distribution highlights the widespread impact of IBD genes across the GIT. Organs where expression data was not available are colored in grey.

**Figure 4:** Expression Patterns of IBD-Associated Genes Across Tissues. **(a)** A heatmap of average expression levels of IBD-associated genes in various regions of the gastrointestinal (GIT) tract, including colon-sigmoid, Colon-transverse, and Small intestine-terminal Ileum. Tissues that we could not find expression data for are colored in grey. **(b)** A heatmap of expression levels of IBD-associated genes in different brain regions, including the Amygdala, Anterior Cingulate Cortex (BA24), Caudate (basal ganglia), Cerebellar Hemisphere, Cerebellum, Cortex, Frontal Cortex (BA9), Hippocampus, Hypothalamus, Nucleus Accumbens (basal ganglia), Putamen (basal ganglia), Spinal Cord (cervical C1), and Substantia Nigra. Brain regions that we could not find expression data for are colored in grey. **(c)** Enriched brain-related KEGG pathways with IBD genes. Significant pathways include the Neurotrophin Signaling Pathway and pathways of neurodegeneration. **(d)** Enriched signaling-related KEGG pathways with IBD genes. Significant pathways include the FoxO Signaling Pathway, Calcium Signaling Pathway, and Notch Signaling Pathway. **(e)** An OncoPrint representation of single nucleotide polymorphisms (SNPs) associated with IBD, CD, and UC.

Figure 4b shows the expression of these genes in different brain regions. Notably, high expression levels are observed in the cerebellum, frontal cortex, and hippocampus, indicating potential links between IBD and neurological. This cross-tissue analysis underscores the systemic nature of IBD and its potential impact beyond the GIT.

To further understand the functional implications of the transcriptomic data, we conducted pathway enrichment analyses. Figure 4c presents brain-related KEGG pathways enriched with IBD genes. Significant pathways include the Neurotrophin Signaling Pathway (10 genes, FDR= 1.9E-4, nFold=5.35), which plays a crucial role in the development and function of neurons (Huang E, Reichardt L, 2001), and the Pathways of Neurodegeneration (17 genes, FDR= 6.0E-3, nFold=2.28), highlighting potential links between IBD and neurodegenerative diseases (Kim J et al., 2023). Additionally, the GABAergic Synapse (6 genes, FDR= 1.1E-2, nFold=4.29) and Cholinergic Synapse (6 genes, FDR= 2.9E-2, nFold=3.38) pathways are enriched, reflecting the involvement of IBD genes in neurotransmitter signaling and synaptic function.

To complement the pathways highlighted in Figure 1b, additional signaling pathways were found to be enriched with IBD-associated genes and presented in Figure 4d, emphasizing the breadth of disturbed signaling networks in IBD. There were 41 enriched KEGG pathways that were related to signaling. The most enriched pathways include the NF-kappa B signaling pathway (nGenes=13, FDR=4.0E-07, nFold=7.96), JAK-STAT signaling pathway (nGenes=14, FDR=6.0E-06, nFold=5.50), HIF-1 signaling pathway (nGenes=11, FDR=1.9E-05, nFold=6.43), TNF signaling pathway (nGenes=11, FDR=2.3E-05, nFold=6.25), and NOD-like receptor signaling pathway (nGenes=13, FDR=7.3E-05, nFold=4.60). These pathways are crucial for mediating immune responses and inflammation in IBD, similar to those observed in UC (Holmqvist E et al., 2010; Choi S et al., 2000; Coskun M et al., 2013).

Importantly, several pathways are directly relevant to brain function and neurological processes. These include the Neurotrophin signaling pathway (10 genes, FDR=1.9E-4, Fold Enrichment=5.35), which is essential for neuron survival, differentiation, and synaptic plasticity (Huang T, Reichardt L, 2001). The Oxytocin signaling pathway (7 genes, FDR=3.3E-2, Fold Enrichment=2.89) is involved in regulating social behavior and emotional responses along with a role in neuropsychiatric disorders, suggesting that psychiatric patients may be more prone to developing IBD (Jin Y et al., 2023; Zois C et al., 2010).

Additionally, the Calcium signaling pathway (9 genes, FDR=3.9E-2, Fold Enrichment=2.39) and the Sphingolipid signaling pathway (6 genes, FDR=3.3E-2, Fold Enrichment=3.21) are crucial for neurotransmission and neuroinflammatory processes, highlighting potential cross-talk between the gut and brain in IBD pathogenesis. The Notch signaling pathway (4 genes, FDR=3.9E-2, Fold Enrichment=4.32) is another key regulator of cell fate decision and has implications in both immune function and neural development (Lasky J, Wu H, 2005). Furthermore, the Wnt signaling pathway (8 genes, FDR=1.7E-2, Fold Enrichment=3.07) is involved in neurodevelopmental processes and synaptic plasticity, underscoring the potential impact of IBD on neurological health and vice versa (Patapoutian A, Reichardt L, 2000). Overall, these findings underscore the complex interplay of the immune, inflammation, and the brain in the pathogenesis of IBD, suggesting avenues for further research into the gut-brain axis in this condition.

Lastly, Figure 4e provides an OncoPrint representation of single nucleotide polymorphisms (SNPs) associated with IBD, CD, and UC. This visualization highlights the distribution and frequency of SNP variants across these conditions. Key variants include IL23R, which is common across all IBD subtypes. Other notable SNPs include RNU1-150P, PIGPC2, and TNFSF15. Importantly, the majority of genes have specific variants reported in CD, specific in UC, and some that are shared, illustrating the genetic heterogeneity within IBD.

### Common genes between IBD and neurological disorders

Expanding on our previous analyses, we examined the overlap between IBD-associated genes and genes implicated in various brain disorders. There were many previous reports of comorbidities between IBD and neurological conditions (Fakhfouri G et al., 2024; Bernstein C et al., 2019). This analysis provides insights into the shared genetic architecture between these conditions.

We first focused on the genetic overlap among IBD, CD, and UC. We identified 210 genes common to IBD and CD and 271 genes shared across all three conditions. Additionally, there are specific overlaps of 112 genes between IBD and UC and 15 genes unique to both CD and UC, underscoring the genetic similarities and distinctions among the IBD subtypes (Waterman M et al., 2011), as shown in Figure 5a.

**Figure 5:** Genetic Overlap Between IBD and Brain Disorders. (**a**) A Venn diagram showing the overlap of genes associated with IBD and schizophrenia (SCZ). (b) A Venn diagram showing the overlap of genes associated with IBD and depression. **(c)** A Venn diagram showing the overlap of genes associated with IBD and autism spectrum disorder (ASD). **(d)** A Venn diagram showing the overlap of genes associated with IBD and attention-deficit/hyperactivity disorder (ADHD). **(e)** A Venn diagram showing the genetic overlap among IBD, CD, and UC. **(f)** Comprehensive Venn diagram illustrating the genetic interplay between IBD, SCZ, depression, ASD, and ADHD. **(g)** An UpSet plot summarizing the intersections of genes among IBD, SCZ, depression, ASD, and ADHD.

We then examined the genetic overlap between IBD and several neurological disorders. The overlap between IBD and depression reveals 94 shared genes, indicating a genetic link between inflammatory processes in the gut and mood disorders (Goodhand J et al., 2012), as shown in Figure 5b. In the case of autism spectrum disorder (ASD), 52 genes overlap with IBD, highlighting potential shared genetic mechanisms related to immune system regulation and neurological development, as depicted in Figure 5c. For attention-deficit/hyperactivity disorder (ADHD), 68 genes are shared with IBD, suggesting possible common pathways in neurodevelopmental and inflammatory processes, as illustrated in Figure 5d. There are also notable overlaps between genes associated with IBD and schizophrenia (SCZ), with 125 genes being common to both conditions. This suggests potential involvement of the immune system and schizophrenia (Sung K et al., 2022), as represented in Figure 5e.

Further extending this analysis to include all the aforementioned brain disorders, a comprehensive Venn diagram (Figure 5f) illustrates the complex genetic interplay between IBD, SCZ, depression, ASD, and ADHD. The center of the diagram reveals a core set of genes common to all conditions, emphasizing the potential central role of these genes in both inflammatory and neurological processes.

Finally, an UpSet plot (Figure 5g) summarizes the intersections of genes among IBD, SCZ, depression, ASD, and ADHD. The plot highlights the number of genes unique to each condition as well as the shared genes across multiple conditions. Notably, the highest overlap is observed between SCZ and depression. After the overlaps in brain disorders, the overlaps between IBD and each of the brain disorders are shown. This representation provides a clear visualization of the genetic overlap and distinct genetic contributions to each condition.

## Discussion

The present study provides a comprehensive analysis of the genetic landscape of Inflammatory Bowel Disease (IBD), focusing on its two main subtypes, CD and UC. By leveraging genome-wide association studies (GWAS) data and integrating findings from the Genotype-Tissue Expression (GTEx) project, we have explored the expression patterns of IBD-associated genes across various tissues, particularly the gastrointestinal (GIT) tract and the brain. Our analyses highlight the systemic nature of IBD and its potential links to neurological and psychiatric disorders.

### Genetic Insights and Pathway Enrichment

Our study identified several essential genes associated with IBD, CD, and UC, with notable overlaps among these conditions. Genes such as NOD2, IL23R, and STAT3 were consistently highlighted across multiple analyses, underscoring their critical roles in IBD pathogenesis. The pathway enrichment analyses revealed significant involvement of immune-related pathways, including the Inflammatory Bowel Disease Pathway, Th17 Cell Differentiation, NF-Kappa B Signaling Pathway, and Cytokine-Cytokine Receptor Interaction. These pathways are well-known for their roles in mediating immune and inflammatory responses, which are central to the pathology of IBD (Atreya I, Atreya R, Neurath M, 2008; Baumgart D, Carding S, 2007). Overall, there were 72 common Kegg pathways among IBD, CD, and UC, which were enriched with IBD genes. Also, there were seven unique pathways to IBD, ten unique pathways to UC, and two pathways unique to CD.

The enrichment of the HIF-1 Signaling Pathway in both CD and UC, as well as the broader IBD category, highlights the role of hypoxia in modulating inflammatory responses and tissue remodeling in the gut (Kerber E et al., 2020). The JAK-STAT Signaling Pathway, significantly enriched in both CD and UC, is crucial for transducing signals from cytokines and growth factors, influencing gene expression and immune cell function (Coskun M et al., 2013; Schindler C et al., 2007).

### Cross-Tissue Expression Patterns

By integrating GTEx data, we were able to examine the expression of IBD-associated genes across different tissues. Our findings demonstrate that these genes are not only highly expressed in the GIT tract but also in various brain regions, such as the cerebellum, frontal cortex, and hippocampus. This cross-tissue expression pattern suggests that IBD genes may have systemic effects, potentially influencing neurological functions.

The identification of brain-related KEGG pathways enriched with IBD genes, including the Neurotrophin Signaling Pathway and pathways of neurodegeneration, suggests potential links between IBD and neurological disorders (Huang E, Reichardt L, 2001; Kim J et al., 2023). These findings are consistent with previous reports of increased prevalence of neurological and psychiatric conditions, such as schizophrenia, depression, and autism spectrum disorder (ASD), among IBD patients (Benros M et al., 2011; Mikocka-Walus A et al., 2016; Bernstein C et al., 2019).

### Genetic Overlap with Brain Disorders

The genetic overlap between IBD and various brain disorders was further elucidated through Venn diagrams and an UpSet plot. Significant overlaps were observed between IBD and schizophrenia, depression, ASD, and ADHD. These findings indicate shared genetic and immunological pathways, suggesting a common underlying mechanism that may contribute to the comorbidity of these conditions (Reardon S, 2014; Mage M et al., 2023; Udoh P et al., 2009; Sedgwick P, 2013).

The identification of core genes common to IBD and multiple brain disorders underscores the importance of understanding the broader systemic implications of IBD. These shared genes may serve as potential therapeutic targets, offering opportunities for developing treatments that address both gastrointestinal and neurological symptoms.

### Strengths and Limitations

One of the strengths of this study is the integration of large-scale GWAS data with comprehensive expression datasets from the GTEx project. This approach provides a robust framework for identifying key genes and pathways involved in IBD and exploring their broader systemic effects. Additionally, the use of multiple visualization techniques, including Venn diagrams, enhances the interpretability of the complex relationships between genes, pathways, and diseases.

However, there are some limitations to consider. The expression data from GTEx represents a cross-sectional snapshot and may not capture dynamic changes in gene expression over time or in response to environmental factors. Moreover, the study primarily focuses on transcriptional regulation, and further research is needed to explore the functional implications of identified genetic variants at the protein level.

### Future Directions

Future research should aim to validate these findings using longitudinal data and explore the functional roles of identified genes and pathways in detail. Integrating additional omics data, such as proteomics and metabolomics, could provide deeper insights into the molecular mechanisms underlying IBD and its associated comorbidities. Furthermore, investigating the impact of environmental factors, such as diet and microbiota, on gene expression and disease progression could help elucidate the complex interplay between genetic and environmental determinants of IBD.

Induced pluripotent stem cells (iPSCs) have revolutionized the study of various neurological disorders such as Autism Spectrum Disorder, Schizophrenia, bipolar disorder, Parkinson’s disease, and more (Rike W, Stern S, 2023; Tripathi M et al., 2023; Hussein Y, Tripathi U, et al., 2023; Stern S, Zhang L, Wang M, et al., 2024; Figueiredo T et al., 2021; Rosh I et al., 2024; Manole A et al., 2023; Tripathi U et al., 2024) by enabling researchers to create patient-specific neurons and brain organoids. These organoids provide an invaluable platform for modeling complex brain functions and behaviors as well as the underlying pathology of diseases. Researchers can explore the genetic, molecular, and cellular mechanisms implicated in various neurological conditions by converting somatic cells from patients into iPSCs and subsequently differentiating them into neural cell types.

Given the shared signaling pathways and cellular interactions between the gut and the central nervous system, integrating iPSC technology into the gut-brain axis research area presents exciting future directions. One promising avenue involves the development of joint gut-brain organoids derived from iPSCs. These organoids would allow researchers to elucidate the interactions between gut microbiota, the enteric nervous system, and central nervous system neurons in the context of IBD. By studying these organoids, scientists could investigate how inflammatory signals from the gut affect neuronal function and vice versa. Such experiments could lead to a deeper understanding of the mechanisms underlying IBD and its associated neurological comorbidities, such as anxiety and depression, often observed in patients with chronic gastrointestinal disorders.

Furthermore, joint gut-brain organoids could be utilized to screen potential therapeutic compounds that simultaneously target both systems, opening new avenues for integrated treatments that address IBD and its neurological impacts. Ultimately, this innovative approach may provide insights into the interconnectedness of these systems, paving the way for more holistic treatment strategies that recognize the importance of the brain-gut axis in inflammatory bowel disease management.

In conclusion, this study advances our understanding of IBD’s genetic and molecular landscape, highlighting essential genes and pathways involved in disease pathogenesis and their potential links to neurological disorders. These findings pave the way for developing targeted therapeutic strategies and improving patient outcomes by addressing the systemic nature of IBD and its associated comorbidities.

## Funding information

The Zuckerman STEM leadership program and Israel Science Foundation grants – 1994/21 and 3252 /21 for Prof. Shani Stern.

## Conflict of interest/Competing interests

The authors declare no competing interests.

## Code availability

Correspondence and requests for code should be addressed to Prof. Shani Stern.

## Data Availability

All data produced in the present study are available upon reasonable request to the authors

## Acknowledgements

The graphical image for this publication was created with Biorender.com and the figures were plotted in MATLAB.

## Author Contributions

U.T. performed the data curation, data analysis, prepared the figures, and wrote the manuscript. Y.S. contributed to drafting the manuscript. I.D. assisted with data curation and analysis. R.N. created the illustrations using Biorender. E.R. helped in data analysis. E.Z. provided subject matter expertise on Inflammatory Bowel Disease (IBD) and contributed to the interpretation of the results. S.S. conceptualized and supervised the study. All authors reviewed and approved the final version of the manuscript for submission.

## Notes

### Competing Interest Statement

The authors have declared no competing interest.

### Author Declarations

The study used online available GWAS data and GTEx data.

### Summary of Updates

Here, we have added the Authors' contribution section in the manuscript. We have also added a new author.

